# Ethnicity-Specific Effects on Cardiac Arrest During the COVID-19 Pandemic: A Two-Year Prospective Evaluation in a North American Community

**DOI:** 10.1101/2022.10.15.22281071

**Authors:** Harpriya S Chugh, Arayik Sargsyan, Kotoka Nakamura, Audrey Uy-Evanado, Bernadine Dizon, Faye L Norby, Christopher Young, Katy Hadduck, Jonathan Jui, Daniel Shepherd, Angelo Salvucci, Sumeet S Chugh, Kyndaron Reinier

## Abstract

**Background:** Out-of-hospital sudden cardiac arrest (SCA) is a major public health problem with mortality >90%, and incidence has increased during the COVID-19 pandemic. Information regarding ethnicity-specific effects on SCA incidence and survival is lacking.

**Methods:** In a prospective, population-based study of Ventura County, CA residents (2020 Pop. 843,843; 44.1% Hispanic), we compared SCA incidence and outcomes during the first two years of the COVID-19 pandemic to the prior four years, overall and by ethnicity (Hispanic vs non-Hispanic).

**Findings:** Of 2,222 OHCA cases identified, 907 occurred during the pandemic (March 2020 - Feb 2022) and 1315 occurred pre-pandemic (March 2016 - Feb 2020). Overall age-standardized annual SCA incidence increased from 38.9/100,000 [95% CI 36.8-41.0] pre-pandemic to 53.8/100,00 [95% CI 50.3 - 57.3, p<0.001] during the pandemic. Among Hispanics, incidence increased by 77%, from 38.2/100,00 [95% CI 33.8-42.5] to 67.7/100,00 [95% CI 59.5- 75.8, p<0.001]. Among non-Hispanics, incidence increased by 26% from 39.4/100,000 [95% CI 36.9-41.9, p<0.001] to 49.8/100,00 [95% CI 45.8-53.8]. SCA incidence rates closely tracked COVID-19 infection rates. During the pandemic, SCA survival was significantly reduced (15.3% to 10.0%, p<0.001) and Hispanics were less likely than non-Hispanics to have bystander CPR (44.6% vs. 54.7%, p=0.005) and shockable rhythm (15.3% vs. 24.1%, p=0.003).

**Interpretation:** Hispanic residents experienced higher SCA rates during the pandemic with less favorable resuscitation profiles. These findings implicate potential ethnicity-specific barriers to acute care and represent an urgent call to action at the community and health-system levels.

**Funding:** National Heart Lung and Blood Institute Grants R01HL145675 and R01HL147358.

## Introduction

Out-of-hospital sudden cardiac arrest (SCA) is a mostly fatal condition with an annual incidence of 40-80 per 100,000, and a US average survival rate of 9%.^1,2^ The COVID-19 pandemic has led to an increase in SCA incidence with a decline in survival from SCA.^3^

The U.S. Hispanic population has experienced a higher incidence of COVID-19 than the Non-Hispanic White population,^4,5^ with disproportionate increases in overall cardiovascular mortality during the pandemic.^6^ However, there is a lack of information regarding pandemic-related changes in SCA incidence among Hispanics. In an earlier study conducted pre-pandemic, we reported that Hispanic and Non-Hispanic White residents of Ventura County, CA had similar age-standardized incidence of SCA.^7^ In the present analysis, we evaluated incidence of and survival from SCA among Hispanic and non-Hispanic individuals in Ventura County, CA during two years of the COVID-19 pandemic.

## Methods

### Setting

Individuals with out-of-hospital SCA from Feb. 1, 2015 through Feb. 28, 2022 were identified from the ongoing population-based PRESTO (PREdiction of Sudden Death in MulTi-Ethnic COmmunities) study in Ventura County, CA.^7^ In this study, all incident cases of presumed SCA requiring cardiopulmonary resuscitation and/or defibrillation by the county’s 2-tiered Emergency Medical Services (EMS) system were prospectively identified and adjudicated by study physicians based on detailed review of the EMS pre-hospital care report, pre-arrest and at-arrest medical records, death certificates, and autopsies when available. Adjudicated SCA was defined as a sudden, unexpected pulseless collapse of likely cardiac origin;^8^ all cases with an identifiable non-cardiac etiology were excluded, such as trauma, drug overdose, and chronic terminal illness (e.g., malignancy not in remission). The PRESTO study was approved by the Institutional Review Boards of the Ventura County Medical Center, Cedars-Sinai Medical Center, and all participating health systems. All survivors of SCA provided informed consent; for non-survivors, this requirement was waived. This report follows the STROBE guidelines for reporting results of observational studies.^9^

### Outcomes

The primary outcome was incidence of SCA. A secondary outcome was survival to hospital discharge (STHD) following SCA.

### Definition of Ethnicity

Race and ethnicity were obtained from death certificates for the majority of cases (87%), and also from medical records or interviews with survivors. Death certificates for defining Hispanic ethnicity have a positive predictive value of 91.3% compared to self-report.^10^ Missing ethnicity (4%) was imputed by surname based on US Census data.^11^ Individuals of Hispanic ethnicity were compared to all other cases.

### Variable definitions

Response time was defined as the time between 911 call and EMS personnel arrival at the patient’s side, bystander cardiopulmonary resuscitation (CPR) was defined as CPR provided by individuals not part of the organized EMS response, shockable rhythm was defined as ventricular fibrillation/ventricular tachycardia measured by EMS responders (vs. other non-shockable rhythms such as pulseless electrical activity or asystole). Survival from SCA was defined as survival to hospital discharge (STHD).

### COVID-19 Data Sources

Counts of new COVID-19 cases per month for Ventura County were obtained from the California Department of Public Health.^12^

### Evaluation of COVID-19 positivity or COVID-related illness in SCA cases

To evaluate whether COVID-19 illness was associated with each SCA case during the pandemic period, we evaluated cause of death data from death certificates and medical examiner notes from autopsy when available. From EMS records, we reviewed the narrative and patient clinical notes from the cardiac arrest. For survivors, we reviewed findings from the medical work-up at the time of cardiac arrest. From each source, we performed a text search for keywords “COVID”, “SARS”, or “corona” followed by manual review of those with the keywords. In addition, we performed a manual medical chart review for all individuals with a visit to a medical provider within one month (31 days) prior to SCA during the pandemic period for evidence of a positive COVID-19 test or COVID-related illness.

### Statistical Analysis

We compared SCA incidence overall and by ethnicity (Hispanic vs. non-Hispanic) during the two-year COVID-19 pandemic period (Mar 1, 2020 – Feb 28, 2022) versus the four-year pre-pandemic period (Mar 1, 2016 – Feb 29, 2020) in Ventura County, California (2020 pop. 843,843). We also compared SCA incidence during pandemic peak and non-peak periods to incidence in corresponding months in the 4 years pre-pandemic. Peak periods were defined as months with ≥3000 new reported COVID-19 cases. Non-peak periods were defined as months with <3,000 new COVID-19 cases reported. In a sensitivity analysis limited to the 2 pandemic years, we compared SCA incidence during COVID peaks vs. non-peak times.

We calculated annualized age-standardized incidence of SCA per 100,000 using the overall and ethnicity-specific population of Ventura County, CA in 2020, with the total US population as the standard (U.S. Census Bureau ACS 2020 tables B01001 and S0101).

We compared univariate demographic and arrest circumstances and outcomes in the pandemic vs. pre-pandemic periods with *t-*tests and chi-square tests, overall and by ethnicity. Adjusted odds ratios for STHD in the pandemic vs pre-pandemic period were calculated in multivariable logistic regression models with terms for age, sex, response time, bystander CPR, shockable rhythm, ethnicity, and pandemic period. An interaction term (ethnicity by pandemic period (vs pre)) was used to test for an ethnicity-specific change in STHD during the pandemic. Analysis followed STROBE guidelines^9^ using SAS Version 9.4 (SAS Institute, Cary, NC) and a 2-sided significance level of 0.05.

## Results

### Overall pandemic vs. pre-pandemic periods

#### SCA case counts and demographics

The two-year pandemic period included 907 SCA cases (453 per year); the four-year pre-pandemic period included 1315 SCA cases (319 per year). During the pandemic period, monthly SCA case counts showed a marked increase as monthly COVID counts increased in the second COVID wave (Nov 2020 – Feb 2021) (Figure 1). During the pandemic, Non-Hispanic White individuals accounted for a smaller proportion of SCA cases while the proportion Hispanic increased from 24.3% to 31.6% (p<0.001), and mean age decreased during the pandemic (Table 1).

**Table 1.** Characteristics of individuals with out-of-hospital sudden cardiac arrest in Ventura County, CA during the 4-year pre-pandemic period (Mar 2016 – Feb 2020) and the 2-year COVID pandemic period (Mar 2020 – Feb 2022).

|  | SCA Cases<br>4 Year Pre-Pandemic<br>Period<br>(n=1315) | SCA Cases<br>2 Year Pandemic<br>period<br>(n=907) | p-value |
| --- | --- | --- | --- |
| Race / ethnicity |  |  | <0.001 |
| White | 851 (64.7%) | 483 (53.3%) |  |
| Black | 25 (1.9%) | 22 (2.4%) |  |
| Asian | 71 (5.4%) | 60 (6.6%) |  |
| Hispanic | 320 (24.3%) | 287 (31.6%) |  |
| Other | 15 (1.1%) | 13 (1.4%) |  |
| Missing | 33 (2.5%) | 42 (4.6%) |  |
| Age (mean ± SD), years | 71.3 ± 15.8 | 69.5 ± 17.0 | 0.01 |
| Male | 857 (65.2%) | 586 (64.6%) | 0.78 |
| *Arrest Circumstances* |  |  |  |
| Witnessed arrest | 637 (48.4%) | 418 (46.3%) | 0.33 |
| Bystander CPR | 742 (56.4%) | 467 (51.5%) | 0.02 |
| Response time >4 min | 1031 (78.6%) | 812 (89.9%) | <0.001 |
| Shockable rhythm | 335 (25.5%) | 193 (21.3%) | 0.02 |
| Survival to hospital discharge | 201 (15.3%) | 90 (10.0%) | <0.001 |

Table presents n (%) except where noted.
Data on witnessed arrest missing for 5 cases during the pandemic period; data on survival to hospital discharge missing for 6 cases during the pandemic period.

**Figure 1:**
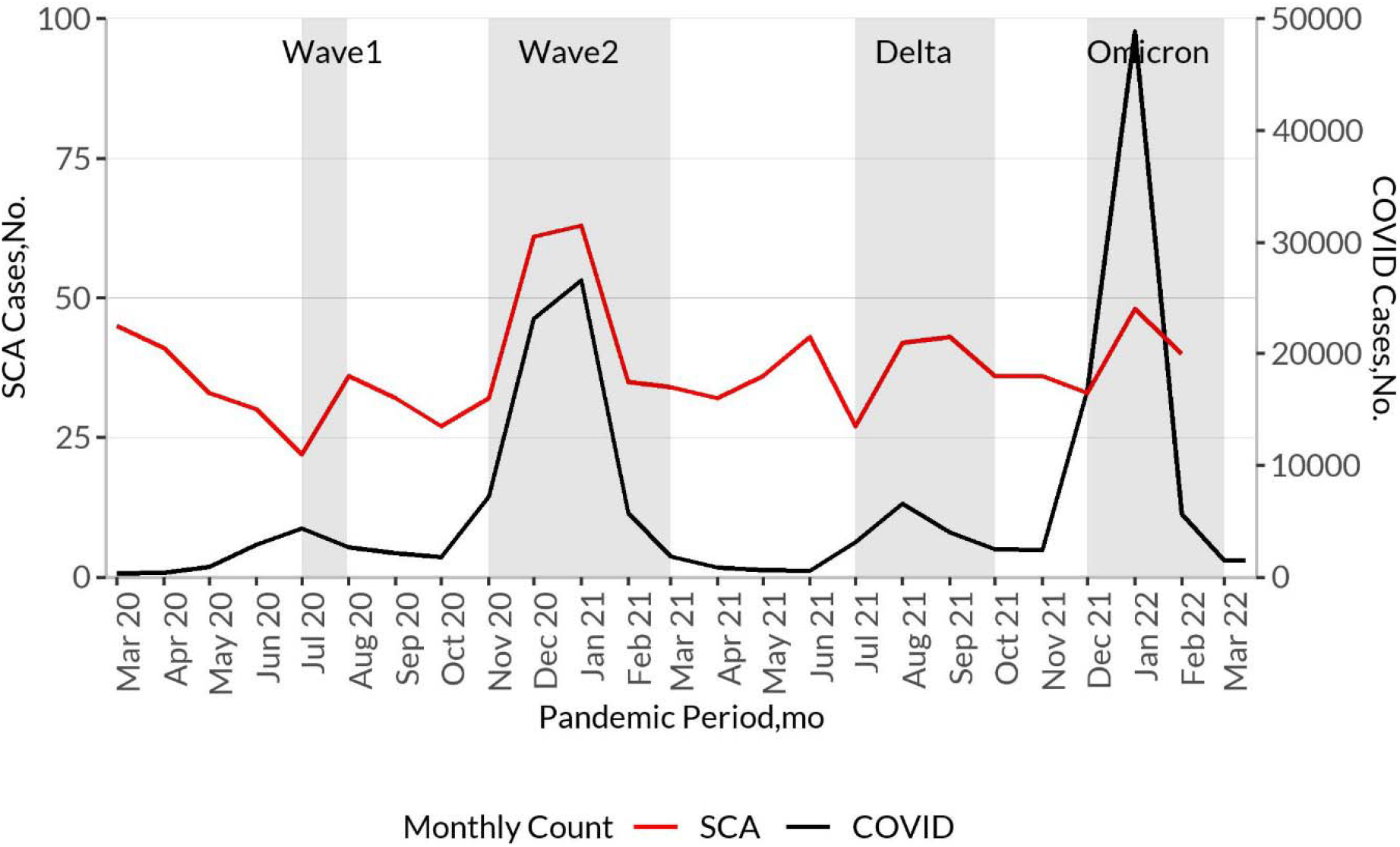
Monthly counts of COVID-19 cases and out-of-hospital sudden cardiac arrest (SCA), Ventura County, CA, pandemic period (March 2020 – Feb 2022). Black line indicates new monthly COVID-19 cases; red line indicates new monthly SCA cases. Grey areas indicate peaks during the pandemic with ≥3000 monthly COVID-19 cases. Four COVID peak periods were observed: 1 month (wave 1, July 2020), 4 months (wave 2, Nov 2020-Feb 2021), 3 months (delta wave, July-Sep 2021), and 3 months (initial omicron wave, Dec 2021-Feb 2022). Non-peak periods were defined as months with <3,000 new COVID-19 cases reported (non-shaded areas).

#### Incidence

Overall age-standardized annual incidence of SCA increased by 38% from 38.9 [95% CI 36.8 – 41.0] per 100,000 before the pandemic to 53.8 [95% CI 50.3 - 57.3] per 100,000 during the pandemic (Table 2). Among Hispanics, age-standardized incidence per 100,000 increased by 77% from 38.2 [95% CI 33.8-42.5] to 67.7 [95% CI 59.5-75.8] (p<0.001, Table 2). Among non-Hispanics, age-standardized incidence per 100,000 increased by 26% from 39.4 [95% CI 36.9-41.9] to 49.8 [95% CI 45.8-53.8](p<0.001, Table 2). Pre-pandemic, SCA incidence did not differ by ethnicity (p=0.62) but during the pandemic, SCA incidence was significantly higher among Hispanics than among non-Hispanics (p<0.001).

**Table 2.** Out-of-hospital cardiac arrest incidence in Ventura County, CA during the COVID-19 pandemic (March 2020 – Feb 2022), overall and during COVID-19 peak and non-peak periods compared to the corresponding pre-pandemic periods (March 2016 – Feb 2020).

|  | Age-standardized incidence per 100,000 [95% CI] |  | Age-standardized mean increase in incidence [95% CI] | p-value |
| --- | --- | --- | --- | --- |
|  | Pre-Pandemic Period | Pandemic Period |  |  |
| *Entire period* | n=1315 | n=907 |  |  |
| All individuals | 38.9 [36.8-41.0] | 53.8 [50.3-57.3] | 14.9 [10.8-19.0] | <0.001 |
| Hispanic | 38.2 [33.8-42.5] | 67.7 [59.5-75.8] | 29.5 [20.3-38.7] | <0.001 |
| Non-Hispanic | 39.4 [36.9-41.9] | 49.8 [45.8-53.8] | 10.4 [5.7-15.0] | <0.001 |
| *Peak Periods\** | N=625 | N=446 |  |  |
| All individuals | 40.3 [37.1-43.4] | 57.8 [52.4-63.1] | 17.5 [11.3-23.7] | <0.001 |
| Hispanic | 45.4 [38.4-52.4] | 77.2 [64.4-90.1] | 31.9 [17.2-46.5] | <0.001 |
| Non-Hispanic | 38.9 [35.3-42.6] | 52.1 [46.1-58.1] | 13.2 [6.1-20.2] | <0.001 |
| *Non-Peak Periods\** | N=690 | N=461 |  |  |
| All individuals | 37.8 [35.0-40.6] | 50.4 [45.8-55.1] | 12.6 [7.2-18.1] | <0.001 |
| Hispanic | 32.0 [26.7-37.4] | 59.5 [49.2-69.9] | 27.5 [15.8-39.2] | <0.001 |
| Non-Hispanic | 39.8 [36.4-43.2] | 47.8 [42.6-53.1] | 8.0 [1.8-14.3] | 0.01 |

\* Peak periods were defined as months with $\geq 3000$ new reported COVID-19 cases (Figure 1, Panel A). Four peaks were observed: 1 month (first wave, July 2020), 4 months (second wave, Nov 2020-Feb 2021), 3 months (delta wave, July-Sep 2021), and 3 months (initial omicron wave, Dec 2021-Feb 2022). Non-peak periods were defined as months with $< 3,000$ new COVID-19 cases reported. For pre-pandemic comparisons, incidence during the combined months of the 4 peak periods was compared to the same months in the pre-pandemic period. For comparisons of non-peak periods, all other months during the pandemic were compared to those same months pre-pandemic.

### COVID peak and non-peak months

#### Incidence compared to corresponding pre-pandemic periods

During the pandemic, Ventura County had four “peaks” of ≥3000 new COVID-19 cases per month, lasting 1 month (first wave, July 2020), 4 months (second wave, Nov 2020-Feb 2021), 3 months (delta wave, July-Sep 2021), and 3 months (initial omicron wave, Dec 2021-Feb 2022) (grey areas, Figure 1). During combined COVID peak months (11 months total with ≥3000 new COVID cases), incidence of SCA increased by 17.5 per 100,000 (p<0.001) compared to the same months in the pre-pandemic period (Table 2). During non-peak periods, incidence of SCA also increased by 12.6 per 100,000 (p<0.001) (Table 2). Among Hispanic individuals, the SCA increase during COVID peak times (31.9 per 100,000) was larger than among non-Hispanic individuals (13.2 per 100,000); Hispanic individuals also had a larger increase in SCA incidence during non-peak times (Hispanic 27.5 per 100,000 and non-Hispanic 8.0 per 100,000) (Table 2). When calculated for each peak and non-peak period separately, SCA incidence increased markedly during the second COVID wave among Hispanic residents (Figure 2, Panel A) and remained higher than pre-pandemic levels throughout the 2-year period, in both COVID peak and COVID non-peak periods; among non-Hispanic residents, a smaller increase in SCA incidence was apparent during the second COVID wave, with smaller differences from pre-pandemic levels throughout the remaining pandemic period (Figure 2, Panel B).

**Figure 2:**
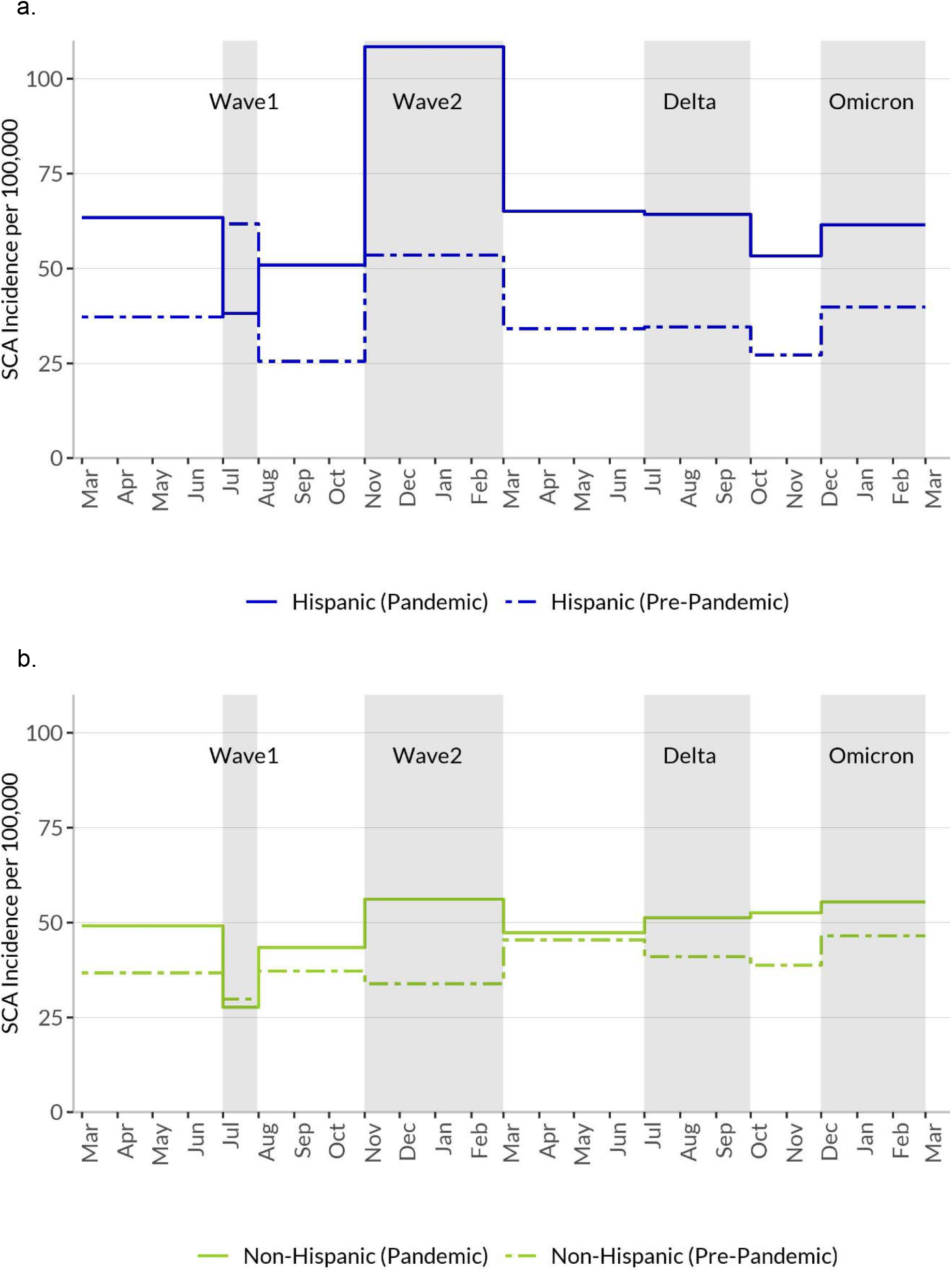
Age-standardized annualized SCA incidence in Ventura County, CA among Hispanics and Non-Hispanics during COVID pandemic peak and non-peak periods (March 2020 – Feb 2022) compared to the same months in the 4-year pre-pandemic period (March 2016 – Feb 2020). <u>Panel A.</u> SCA incidence among Hispanics during each peak and non-peak period. Incidence during pandemic periods indicated by solid blue lines; pre-pandemic periods by dashed blue lines. A pronounced increase in SCA incidence was observed in the second COVID wave (Nov 2020 – Feb 2021), and incidence remained higher than pre-pandemic levels in subsequent peak and non-peak periods. <u>Panel B.</u> SCA incidence among Non-Hispanics. Incidence during pandemic periods indicated by solid green lines; pre-pandemic periods by dashed green lines. A smaller increase in SCA incidence was observed and incidence remained higher, with less apparent pandemic differences than among Hispanics.

#### Incidence within pandemic period

Within the 2-year pandemic period, combined peak periods (11 months) had a higher age-standardized annualized SCA incidence at 57.8 per 100,000 [95% CI 52.4-63.1] than combined non-peak periods (13 months) at 50.4 per 100,000 [95% CI 45.8-55.1] (p=0.04). Among Hispanics, SCA incidence increased from 59.5 [49.2-53.1] to 77.2 [64.4-90.1] (p=0.04) during non-peak vs. peak periods. Among Non-Hispanics, SCA incidence increased from 47.8 [42.6-53.1] to 52.1 [46.1-58.1] (p=0.30). The peaks in SCA incidence in the later COVID surges were less evident (Figure 1 and Figure 2).

#### Resuscitation circumstances and survival from SCA

Compared to the pre-pandemic period, EMS response times were prolonged, bystander CPR decreased (from 56.4% to 51.5%, p=0.02), and shockable rhythm declined (from 25.5% to 21.3%, p=0.02) (Table 1). Survival to hospital discharge (STHD) declined significantly during the pandemic from 15.3% to 10.0% (p <0.001, Table 1 and Figure 3).

**Figure 3:**
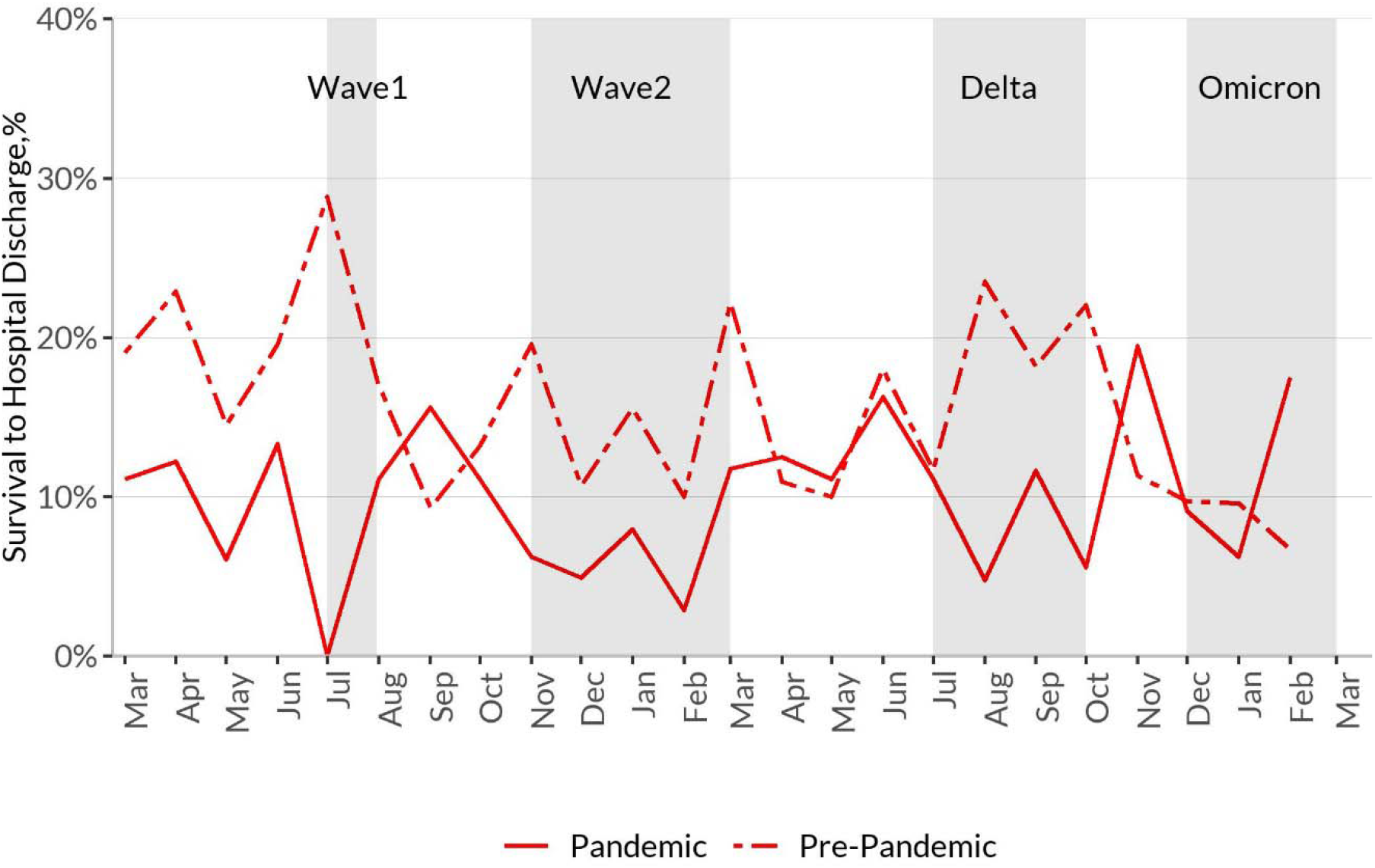
Survival to hospital discharge by month, pandemic vs. pre-pandemic. Pandemic indicated by solid red line; pre-pandemic indicated by dashed red line. Monthly percent survival to hospital discharge (STHD) compared to the average monthly percent STHD during the 4-year pre-pandemic period. STHD was below pre-pandemic levels during most months of the pandemic, particularly in the waves 1 and 2, and the Delta wave.

During the pre-pandemic period, the only apparent ethnicity-specific difference in arrest circumstances was a smaller proportion of Hispanic individuals with an EMS response time >4 minutes (Figure 4, Panel A). Within the pandemic period, Hispanic cases were less likely than non-Hispanic cases to have bystander CPR (44.6% vs. 54.7%, p=0.005) and shockable rhythm (15.3% vs. 24.1%, p=0.003) (Figure 4, Panel B); these differences were not observed in the pre-pandemic period. When adjusted for demographic and resuscitation factors, STHD remained significantly lower during the pandemic period (adjusted odds ratio 0.63 [95% CI 0.46-0.86] p=0.003), with no significant interaction by ethnicity (p=0.68). Among survivors, cerebral performance scores (CPC) were similar during the two periods (p=0.48).

**Figure 4:**
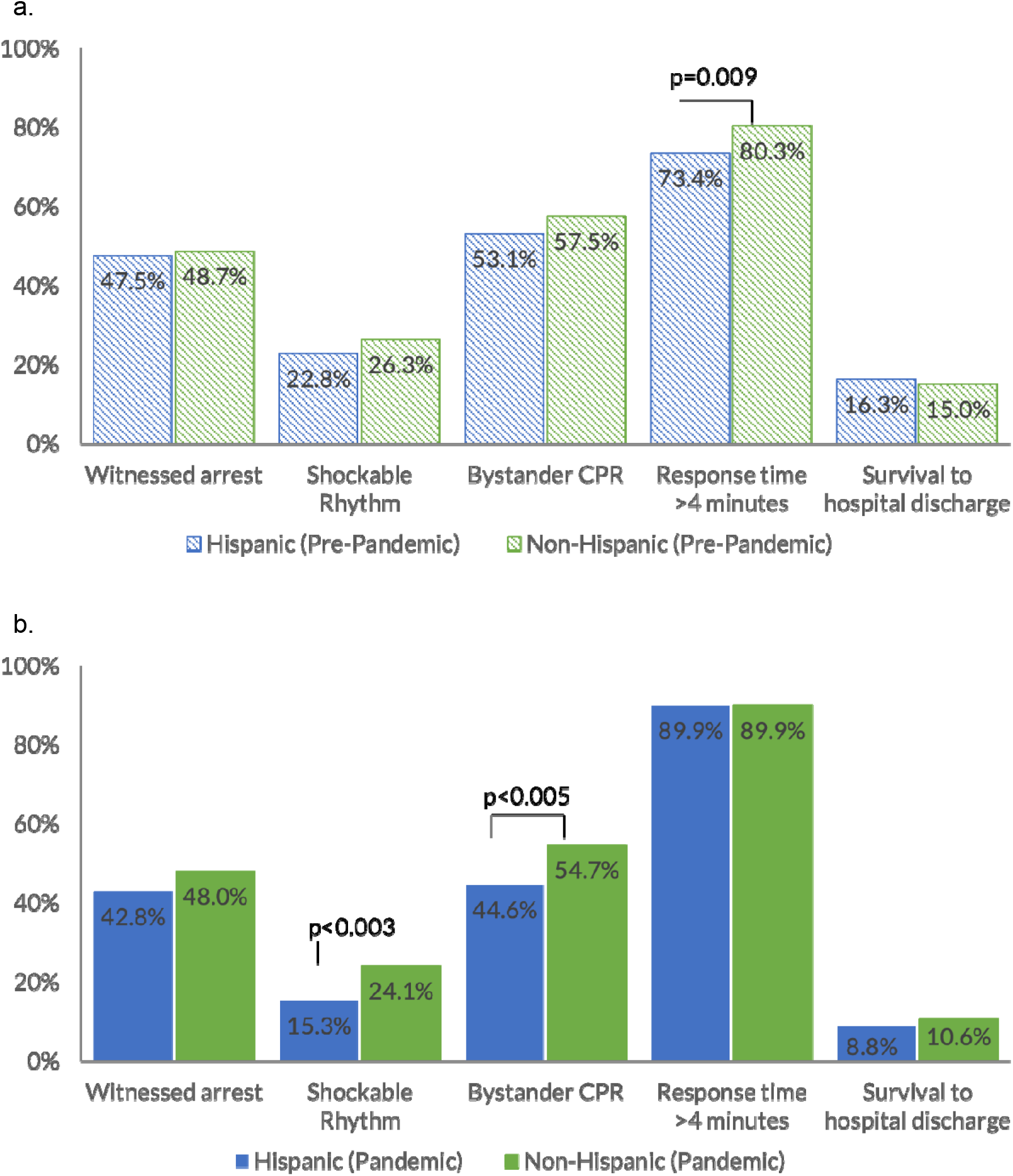
Resuscitation circumstances during pandemic period (March 2020 – Feb 2022) and pre-pandemic period (March 2016 – Feb 2020), Hispanics compared to Non-Hispanics. <u>Panel A:</u> Ethnicity-specific resuscitation circumstances during pre-pandemic period (March 2016 – Feb 2020). Hispanics shown in blue dashed bars; non-Hispanics shown in green dashed bars. A smaller proportion of Hispanics as compared to non-Hispanics had an EMS response time of >4 minutes. <u>Panel B:</u> Ethnicity-specific resuscitation circumstances during pandemic period (March 2020 – Feb 2022). Hispanics shown in blue solid bars; Non-Hispanics shown in green solid bars. Hispanics were less likely than Non-Hispanics to have bystander CPR and shockable rhythm.

### Potential role of acute COVID-19 illness as a factor in SCA

Sixty-two (6.8%) of the 907 SCA cases during the pandemic period had tested positive for SARS-COV-2 or had COVID-19 illness at the time of SCA. Of the 62 SCAs associated with COVID, 43 (69%) were Hispanic. Of the 62 COVID-associated cases, 52 had COVID-19 mentioned on their death certificates (available for 785 (97%) of 811 deceased cases), an additional three were noted at autopsy (available for 132 (16%) of 811 deceased), an additional five were noted as having a positive COVID test or COVID illness in the EMS report from the time of arrest (available for all 907), and an additional two had COVID-19 illness or test positivity mentioned in their medical work-up following SCA (available for 36 (40%) of 90 survivors).

We also reviewed medical charts for 168 of the 907 SCA cases with a medical visit in the 31 days prior to SCA; of these, 55 were inpatient hospital stays. Of the 55, seven individuals (two Hispanic) had COVID-related acute illness (pneumonia (n=5), pneumonia with elevated troponin, ischemia, and sepsis (n=1), and hypoxic respiratory failure (n=1)); and one inpatient had an acute MI ten days before SCA and tested positive for SARS-COV-2 with no acute COVID illness. One additional individual tested positive for SARS-COV-2 but did not have an inpatient visit or COVID-related illness documented, bringing the total to nine individuals with evidence of COVID infection in the month before SCA. Of these nine individuals, five were also identified as COVID-associated in the other records noted above; four had not been identified through our search of death certificates, autopsies, EMS records, and at-arrest medical records, indicating that they were no longer positive or were not evaluated for COVID positivity at the time of their SCA. Of note, all individuals with a hospitalization in the 31 days prior to SCA had to have been discharged from the hospital before their SCA to have been included in our out-of-hospital SCA case group.

## Discussion

Out of hospital SCA incidence increased significantly during the COVID-19 pandemic period, with a larger increase among Hispanic individuals (77% higher incidence than pre-pandemic levels) compared to non-Hispanic individuals (26% higher incidence than pre-pandemic levels). Coinciding COVID-19 and SCA peaks were observed within the pandemic period, particularly during the second COVID wave (winter 2020-2021). A significant decline in STHD was observed during the pandemic, from 15.3% pre-pandemic to 10.0%, with no difference by ethnicity. However, reductions in bystander CPR and shockable rhythm were only observed among Hispanics during the pandemic.

Our results are consistent with literature illustrating two trends that could explain higher SCA incidence among Hispanics during the pandemic: (1) an increase in overall SCA incidence during the COVID pandemic; and (2) higher COVID positivity and higher overall mortality from COVID among Hispanics. Regarding pandemic-era increases in out of hospital cardiac arrest (OHCA), EMS systems from 50 large U.S. cities reported an increase in OHCA of >20% (a few as high as 200%) during early COVID surges, with the timing of increases in OHCA paralleling COVID-19 surges in those cities.^13^ Evidence from individual cities (e.g., Los Angeles, CA^14^ and Paris, France^15^) suggests that increases in SCA were largely due to indirect effects of the pandemic, such as stressors on the EMS system during pandemic surges. Our finding of larger peaks in SCA incidence during COVID case surges is consistent with the EMS 50-cities data.^13^ Multiple factors may also contribute to the association of SCA and COVID-19 peaks, including delays in seeking care among individuals with cardiac symptoms^16^ and delayed EMS activation/response during COVID-19 waves.^17^

Few studies of OHCA/SCA during the pandemic have included data on COVID-19 test positivity or acute illness. One such study reported that of 537 OHCAs from Feb 26 through April 15, 2020 in King County, WA (Seattle area), 6.5% of EMS-treated OHCA had a positive PCR test for SARS-COV-2 or a COVID-like illness (febrile or respiratory illness) based on review of medical records and death certificates.^18^ Our report of 6.8% COVID positivity or COVID illness among the 907 SCA cases during the pandemic was similar, and indicates that a portion of the excess SCA may be due, at least in part, to COVID infection.

Regarding the burden of COVID among Hispanics, there is ample evidence that Hispanics have fared worse than Non-Hispanic Whites in the U.S. due to a variety of factors including systemic issues that disproportionally increase risk or severity of COVID-19 infection among Hispanics, such as working in an essential industry, working while ill, and being more likely to have close contact with COVID-19 affected individuals,^19^ as well as having differential health care access.^20^ A meta-analysis of >4 million patients from 68 studies found that Hispanic individuals were >4.68 times more likely than White individuals to test positive for COVID-19.^5^ Among >200,000 individuals in a Pennsylvania hospital system, Hispanic individuals had higher test positive rates and hospitalizations.^21^ In an analysis of statewide death certificates in California from Feb through July 2020, age-adjusted mortality rate ratios for death from COVID-19 were >4 times higher for Hispanic than for non-Hispanic individuals.^22^ Because COVID-19 infection increases risk for thrombosis, a higher infection rate among Hispanic individuals could translate to a higher event rate for conditions due to thrombosis. Evidence from England and Wales using record-linkage of COVID-19 tests and vascular events showed an adjusted hazard ratio (HR) of 22.1 for myocardial infarction in the first week following COVID-19 diagnosis vs no COVID-19 diagnosis, declining to 4.31 in the second week, but remaining elevated (HR 1.75) through weeks 27-49.^23^ Notably, the HR for myocardial infarction during the first week was as high among non-hospitalized COVID-19 positive individuals as in individuals hospitalized due to COVID, indicating the potential for large population-level effects. While Hispanic individuals accounted for 31.6% of SCA cases during the pandemic, among the 6.8% SCA cases with positive SARS-COV-2 test or COVID-19 illness at the time of SCA, 69% were Hispanic.

Few prior studies have provided direct evidence of a differential impact of the pandemic on SCA by race or ethnicity, and the few that do were conducted early in the pandemic. In New York City from March 1 – April 25, 2020, SCA cases attended by EMS were twice as likely to be Hispanic compared to the year before.^24^

Our findings of less favorable resuscitation characteristics during the pandemic among Hispanics in Ventura County are a cause for concern. In a pandemic setting, community participation in bystander CPR likely drops due to fear of contracting infection and reductions have been reported previously.^17^ Bystander CPR is an established predictor of successful resuscitation leading to a doubling of survival from SCA,^25^ and studies in some communities have reported lower bystander CPR rates in neighborhoods with a higher proportion of Hispanic residents.^26^ In our study population, pre-pandemic differences in bystander CPR were not evident, suggesting the pandemic may have introduced disparities. Presentation with a shockable rhythm is a critical determinant of survival following resuscitation.^25^ Lower rates of shockable rhythms among Hispanics are likely to have more complex explanations. Hispanics may be more likely to delay contact with the health system^27^ for COVID as well as non-COVID illnesses such as acute coronary syndromes, potentially leading to presentation to EMS providers with more critical illness. Ethnicity-specific differences in baseline clinical profile such as obesity, diabetes and renal disease may also contribute.^7^ However, this is the first community-based study to report ethnicity-specific reductions in bystander CPR and shockable rhythms during the pandemic. There could be significant opportunities to improve resuscitation characteristics among Hispanics by targeted awareness and education approaches at the community level.

### Strengths and Limitations

Strengths of our study include data from the first two years of the pandemic (previous studies have been limited to a few months to one year), with detailed analysis of differences by ethnicity, and evaluation of the timing of SCA relative to COVID surges. It is also one of few studies with information regarding individual COVID status and SCA. However, there are some limitations to consider. Due to the lethality of SCA, most ethnicity data were obtained from death certificates rather than self-report. However, this method has been shown to have a positive predictive value of >90% for identifying Hispanic individuals relative to self-report,^10^ and errors in death certificate-determined ethnicity tend to under-ascertain Hispanic individuals.^28^ Therefore, because death certificate-determined ethnicity errors would tend to misclassify Hispanics as non-Hispanics, our results regarding the impact of the pandemic on SCA among Hispanics are likely an underestimation.

We reviewed multiple sources of information (death certificates, autopsies, EMS records at arrest, post-arrest medical work-up, and medical visits from the month prior to SCA) and reported that 6.8% of the 907 SCAs during the pandemic were among individuals whose records indicated a positive SARS-COV-2 test or acute COVID-19 illness, with an additional 4 (0.4%) having a positive SARS-COV-2 test or acute COVID-19 illness in the month before SCA. Because many individuals with SCA die in the field and the majority do not undergo autopsy or additional testing, and because home tests became common in the latter half of our study period and results are not usually available, we believe the number of SCAs occurring in COVID-19 positive individuals is likely an underestimate.

Finally, our data on SCA are limited to individuals with attempted resuscitation, and therefore cannot be generalized to the entire population of out of hospital cardiac arrest (OHCA) including individuals who are dead when EMS arrives, who can make up a substantial proportion of cases;^18^ additionally, the proportion of OHCA who were dead on arrival increased during the early pandemic period.^14^

## Conclusion

The first two years of the COVID-19 pandemic have led to persistently higher SCA incidence, with disproportionately higher increases among Hispanics in this population. Survival from SCA has also decreased with less favorable resuscitation profiles in Hispanics. These findings implicate potential ethnicity-specific barriers for access to acute care during the pandemic and represent an urgent call to action at the community as well as health-system levels.

## Data Availability

Deidentified participant data will be made available after publication upon reasonable request to the corresponding author, following approval of a proposal and a signed data use agreement.

